# U-shaped Association Between Mean Platelet Volume And Short-term Survival In Chinese Patients With Heart Failure

**DOI:** 10.1101/2023.06.16.23291510

**Authors:** Yuan-lei Huang, Qi Zhou, Tao Zheng

## Abstract

**Background:** Mean Platelet Volume (MPV) has been proposed as a potential predictor of increased mortality risk at 6 months among Chinese patients with heart failure (HF). However, the current evidence supporting this association is limited.

**Methods:** This study aimed to investigate the relationship between MPV and HF short-term survival status. The data was obtained from a publicly accessible HF database in Zigong, Sichuan, and included information on 2008 Chinese patients. Baseline MPV was considered as the exposure while HF short-term survival status was the outcome. Two models, a binary logistic regression model and a two-piecewise linear model, were used to analyze the data.

**Results:** The study revealed a U-shaped relationship between MPV and all-cause mortality in HF patients. When MPV levels were less than 9.8, every unit increase in MPV was associated with a 91% reduction (RR: 0.09; 95% CI: 0.03-0.24; P=0.0001) in the risk of death over the next six months. In contrast, at MPV levels above 9.8, each unit increase in MPV was linked to a 27% increase (RR: 1.27; 95% CI: 1.01-1.61; P=0.0434) in the probability of dying within the same period. Stratification by obesity status revealed no significant association between MPV and death in the obese population, while the same U-shaped association was observed among non-obese participants.

**Conclusion:** The present study provides evidence of a U-shaped association between MPV and short-term survival in Chinese patients with heart failure. These findings suggest that MPV may serve as a potential prognostic marker for HF. However, further studies are needed to validate these results and to explore the underlying mechanisms of this association. The observed U-shaped association did not apply to obese patients, suggesting that the effect of MPV on mortality risk in HF patients may be influenced by body weight.

## Introduction

The mean platelet volume (MPV) is a low-cost and easy-to-use blood count indicator that has been suggested to play a critical role in platelet reactivity, inflammation, thrombosis, and heart failure (HF) in previous studies(1-4). MPV is frequently used as a surrogate measure of platelet function since it reflects both platelet size and activity, with larger platelets being associated with increased platelet aggregation, thromboxane production, platelet globulin release, and adhesion molecule expression(5), Elevated MPV levels have been found in various disease states, particularly in individuals with cardiovascular risk factors such as smoking, diabetes, obesity, hypertension, and hyperlipidemia (6-10), indicating a need for careful cardiovascular risk assessment (11). Excessively high MPV levels are also associated with increased cardiovascular disease risk and a poor prognosis.

HF is a worldwide pandemic disease that affects at least 26 million people worldwide and is increasing in prevalence as the population ages(12). In China, the number of people suffering from HF has risen to 1.3 percent of the entire population(13). HF not only affects patients’ quality of life, but it also has a substantial financial impact on their families and society. According to figures, HF accounts for 17 percent of total health expenditures for cardiovascular illness in the United States(14), and total expenses are expected to rise by 127 percent between 2012 and 2030(15). Given the high mortality rate as well as the high financial burden caused by HF, researchers have called HF “the last war of cardiovascular disease in the 21st century.”(16, 17)

In recent years, MPV has piqued interest as a novel prognostic marker in patients with cardiovascular illness. However, evidence linking MPV to all-cause mortality in HF is lacking. Budak et al. discovered a positive correlation between BNP and MPV(18). Furthermore, Kaya (19) and Kandis(2) reported that MPV upon admission was highly linked to in-hospital mortality or death at 6 months, even after controlling for typical HF-related risk variables. However, the majority of the information presented above has come from Western countries. Besides, evidence on the link between MPV and short-term mortality in patients with HF in the Chinese population is limited. Given China’s vast population and rising HF prevalence, a better understanding of the relationship between MPV and short-term mortality risk in HF will be beneficial for future predictive model-driven investigations. In light of this, this study offers a secondary data analysis of a large sample HF database to investigate the relationship between MPV and HF short-term prognosis. This study will also reveal these results.

## Participants and methods

### Data source and study design

This study is a retrospective cohort analysis based on publicly available and published data obtained from clinical records of hospitalized inpatients with confirmed heart failure (HF) at the Fourth People’s Hospital in Zigong City, Sichuan Province, China, between December 2016 and June 2019. The data was downloaded from the PhysioNet website (10.13026/8a9e-w734), and a detailed description of the dataset is available in the literature(20). The dataset has been de-identified to protect patient privacy. All data were extracted from the hospital’s electronic medical record system, and the Ethics Committee of the Fourth People’s Hospital of Zigong approved the study (approval number: 2020-010). Patient informed consent was not required due to the retrospective nature of the study and the anonymization of patient information. The outcome variable was 6-month all-cause mortality in HF patients, recorded as a dichotomous variable in the original data, and the target independent variable was baseline MPV, recorded as a continuous variable.

### Study population

A sample of 2,008 individuals with HF was initially included in the dataset; however, after applying a set of screening criteria, 103 cases were excluded due to missing MPV data, leaving 1,905 cases for data analysis (for details, refer to Figure 1). The diagnosis of HF was made based on the criteria established by the European Society of Cardiology (ESC), and ICD codes were used to collect data on heart failure. The data provider has furnished specific details on the dataset.

**Figure 1.**
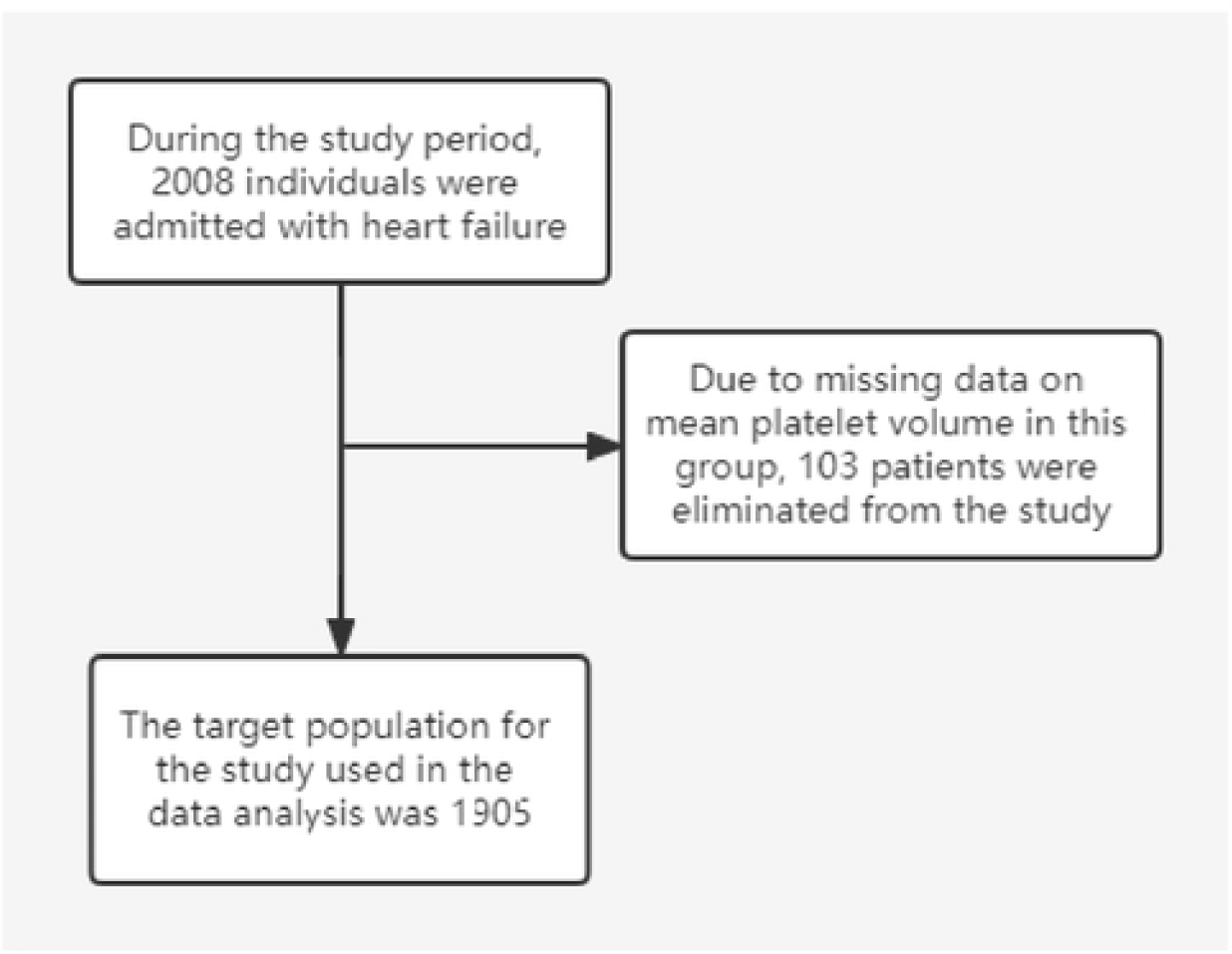

### Variables

#### Mean platelet volume

The mean platelet volume (MPV) was initially recorded as a continuous variable in the raw data. However, to investigate the stability of the results and ensure the robustness of the findings, we further transformed the MPV into a categorical variable based on quartiles. Specifically, the data was divided into four subgroups: Q1 (MPV range: 7.70-10.80, n=475), Q2 (MPV range: 10.90-12.00, n=475), Q3 (MPV range: 12.10-13.20, n=476), and Q4 (MPV range: 13.30-17.60, n=479). This approach allowed us to examine whether the effect values varied equidistantly and to compare the consistency of the results obtained using MPV as a continuous versus a categorical variable.

### 6-month all-cause death in HF patients

The primary outcome measure of this study was six-month all-cause mortality in HF patients. The original dataset provided information on the survival of HF patients at 6 months, represented by a binary variable where 0 indicates survival and 1 indicates death.

### Covariates

In this analysis, potential confounding variables were selected based on prior literature (21-23) that investigated risk factors associated with adverse outcomes in HF, as well as our clinical experience. The following variables were included: age and gender as demographic factors, Charlson Comorbidity Index (CCI) as a measure of co-morbidities, statins, vasodilators, Ras blockers, and positive inotropes as pharmaceutical factors, high-sensitivity troponin, glomerular filtration rate, and BNP as biomarkers, and BMI, NYHY classification, and number of hospitalizations as illness features and anthropometric factors.

### Statistical analysis

Continuous variables were expressed as mean ± standard deviation (SD) (Gaussian distribution) or median (range) (skewed distribution), and categorical variables were expressed as numbers and percentages. The χ2 (categorical variables), one-way ANOVA test (normal distribution), or Kruskal-Whallis H test (skewed distribution) were used to detect differences in the distribution of baseline information across MPV subgroups (quartiles). We examined univariate and multivariate binary logistic regression models to determine the association between the two. To observe the stability of changes in effect values under multiple strategy alterations, we simultaneously display three models. The unadjusted model, or Model 1, does not account for covariates. The model with the fewest adjustments simply for sociodemographic components is Model 2. Model 3 was the one that was completely factor-adjusted. because the binary logistic regression model was unable to account for the nonlinear relationships. We utilized a generalized additive model (GAM) and a smoothed curve fit to examine whether MPV was nonlinearly related to the probability of short-term death in HF (penalized spline method). If nonlinearity was detected, we first used the recursive algorithm + bootstrapping to calculate the inflection point extremely credible intervals, and then constructed two-piece binary logistic regression models on both sides of the inflection point.

To illustrate the robustness of our results, we performed a sensitivity analysis. We transformed mean platelet volume into a categorical variable based on quartiles and calculated p for trend. The aim was to see whether the association with outcome was consistent when MPV was used as a continuous variable versus when it was used as a categorical variable.

R (http://www.r-project.org, R Foundation) and EmpowerStats (http://www.empowerstats.com, X & Y Solutions, Inc., Boston, MA) are statistical applications that were utilized to model the data. Statistical significance was defined as p-values of 0.05 (two-sided).

## Results

### Baseline characteristics of participants

Table 1 summarizes the baseline characteristics of all HF patients, categorized into quartiles based on their MPV levels. Nearly 70% of the patients were below the age of 70. Significant differences were observed between the various MPV level groups (Q1-Q4) for eGFR, Ln BNP, age, sex, use of Ras-Blockers, Inotropes, and diuretics (P < 0.05). Compared to the Q4 group, the Q1, Q2, and Q3 groups had lower eGFR, Ln hs-Tnl, and LnBNP levels, and a smaller percentage of females, a greater percentage of individuals over 70, and a lower percentage of RAS blockers, vasodilators, and diuretics. However, there was no statistically significant difference between the Q1-Q4 groups in terms of BMI, Charlson comorbidity score, NYHA classification, Vasodilator use, Statin use, number of hospitalizations, or 6-month all-cause mortality (P > 0.05).

**Table 1.**
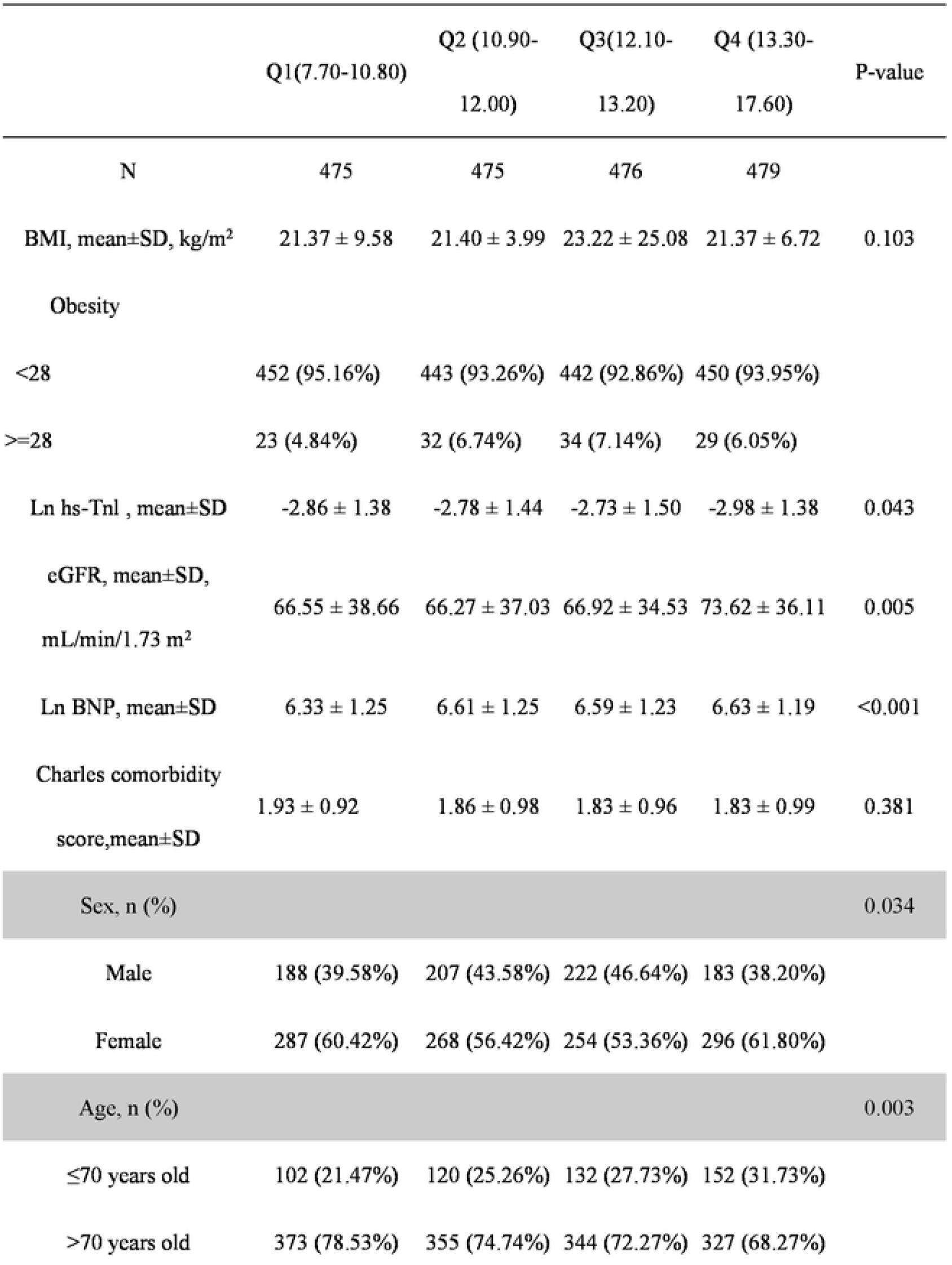
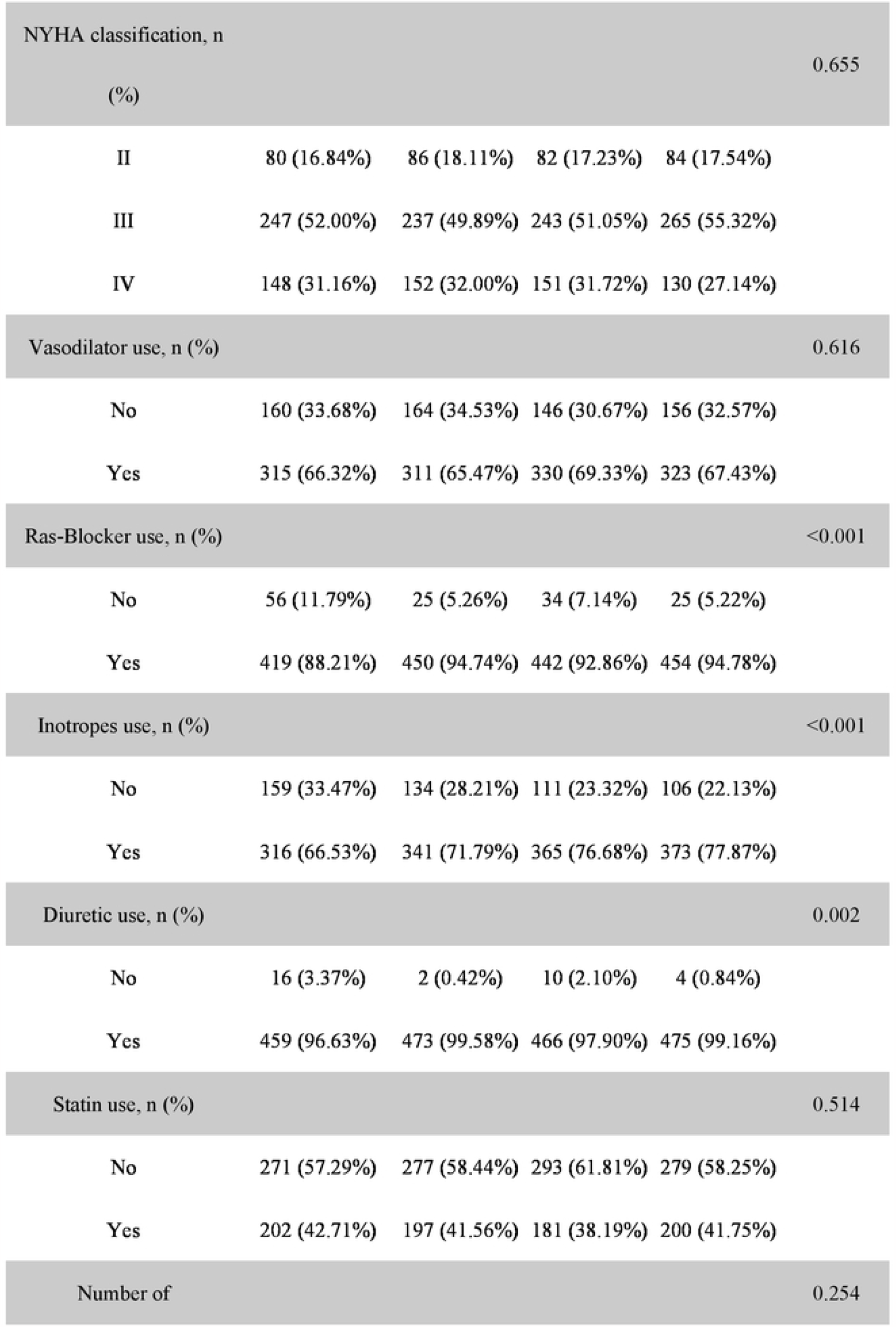
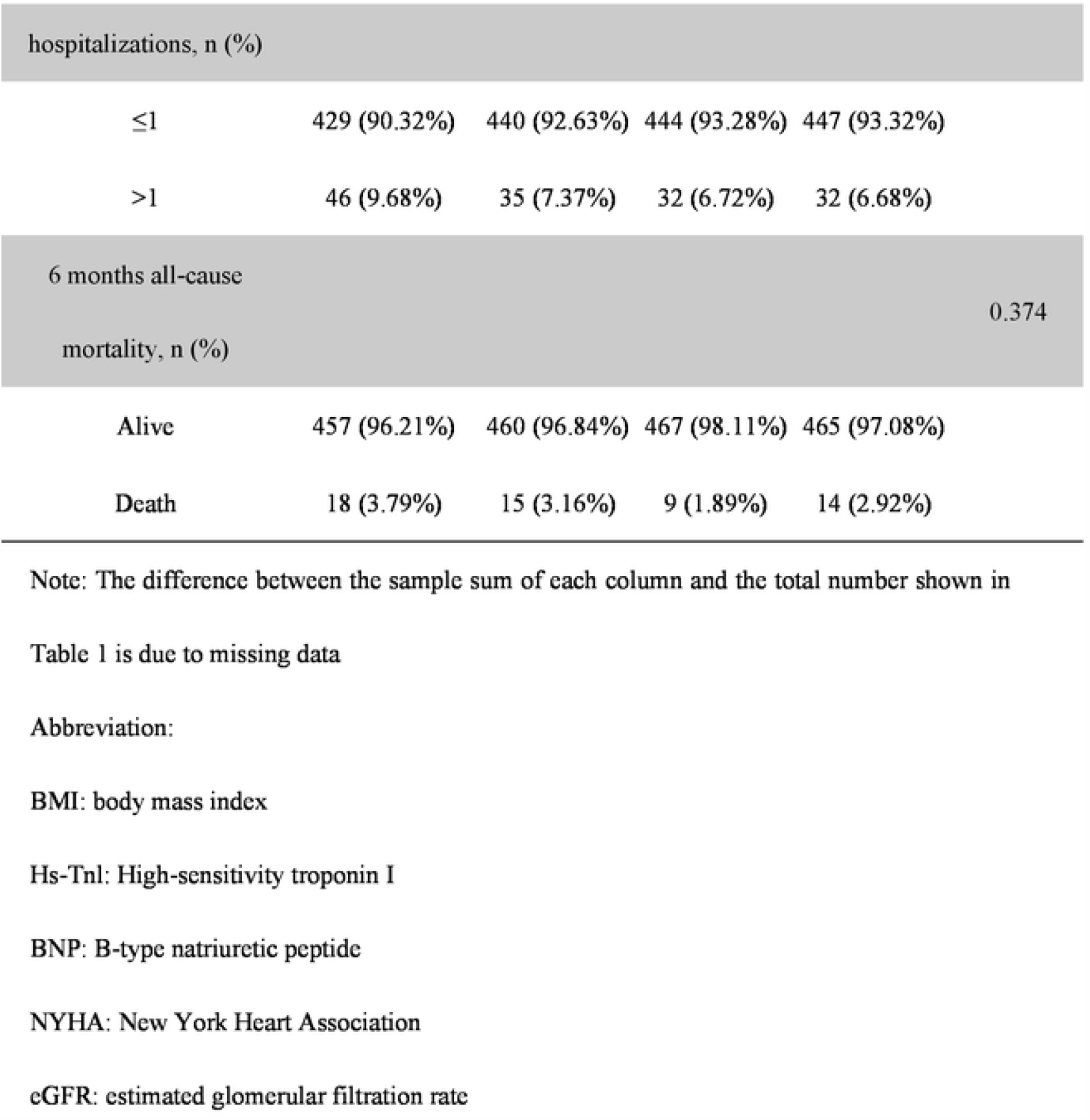
Baseline characteristics of patients with HF grouped by mean platelet volume(quartile)

### Results of a binary logistic regression model’s multivariate analysis

We employed a binary logistic regression model to examine the association between MPV and the 6-month risk of death in HF patients. Our analysis did not reveal a significant relationship between MPV and all-cause mortality (RR, 0.90; 95% CI: 0.77 to 1.06; P=0.1920). Furthermore, the minimally-adjusted (RR, 0.90, 95% CI: 0.77 to 1.06; P=0.2054) and fully-adjusted (RR, 0.94, 95% CI: 0.78 to 1.13; P=0.4888) models yielded identical results despite different adjustment procedures.

To verify the consistency of our findings, we categorized MPV into quartiles and calculated P for trend to compare with the results obtained from MPV as a continuous variable. As shown in Table 2, Our analysis revealed that RR had unequal variation across MPV subgroups (Q1-Q4), and P for trend values were discordant with those obtained using MPV as a continuous variable. These findings suggest that a non-linear relationship between MPV and all-cause mortality cannot be excluded.

**Table 2:**
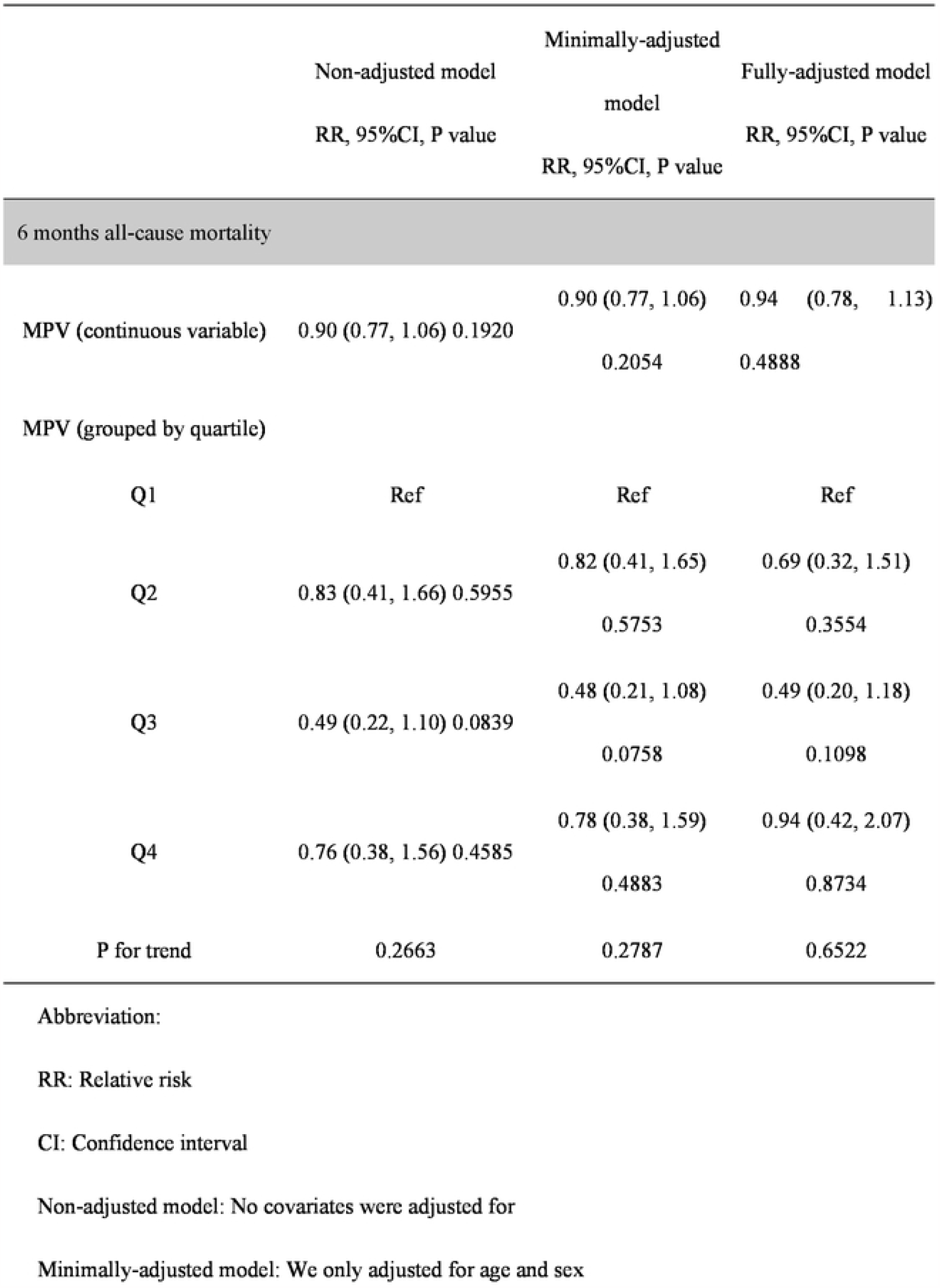
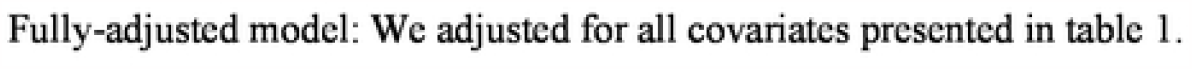
The association between MPV and all-cause mortality using binary logistic regression model

### Using generalized summation models to solve nonlinear issues

We found that the association between MPV and short-term all-cause mortality in HF patients was non-linear using a generalized summation model and smoothed curve fitting (Figure 2). A two-piecewise linear model was developed on both sides of the inflection point, which was obtained using a recursive approach (Table 3). The non-linear results showed a U-shaped relationship between MPV and 6-month mortality. Positive results were observed when MPV was greater than 9.8 (RR, 1.27; 95% CI, 1.01 to 1.61), indicating that each 1fL increase in MPV was associated with a 27% increase in the risk of death. Negative results were found when MPV was less than 9.8 (RR, 0.09; 95% CI, 0.03 to 0.24), indicating that each 1fL increase in MPV was associated with a 91% reduction in the risk of death. Additionally, we stratified the individuals into different obesity categories and found no association between MPV and mortality in the obese group. In contrast, in the non-obese population, the same U-shaped relationship between MPV and 6-month mortality was observed. At MPV<9.7, the association between MPV and 6-month mortality was negative (RR, 0.08; 95% CI, 0.03 to 0.22), indicating that each 1fL increase in MPV was associated with a 92% reduction in mortality risk; at MPV > 9.7 1fL, the association between MPV and 6-month mortality was positive (RR, 1.29; 95% CI, 1.02 to 1.64), indicating that each 1fL increase in MPV was associated with a 29% increase in the risk of death.

**Table 3:**
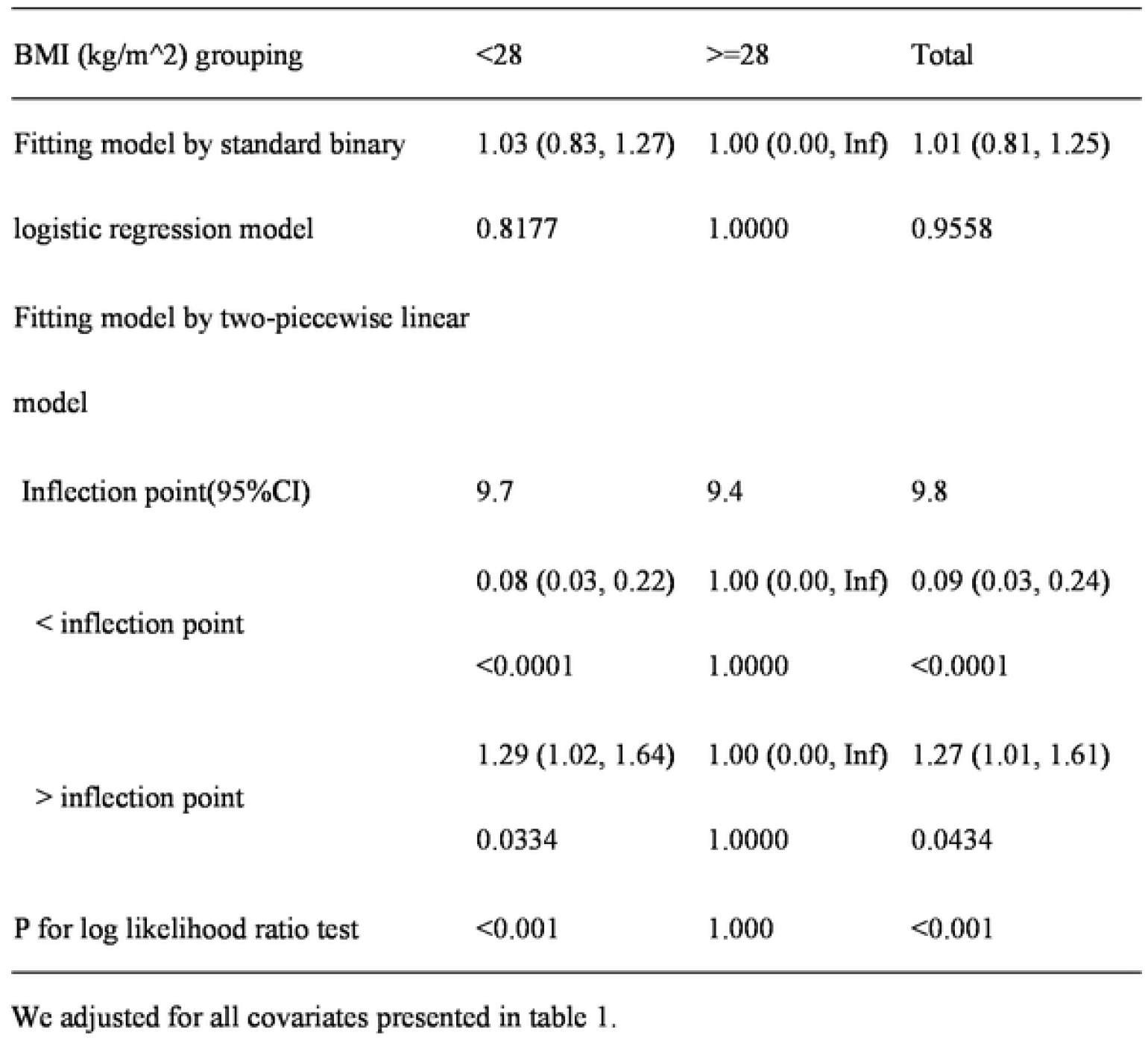
Exploring the non-linear relationship between MPV and all-cause mortality

**Figure 2.**
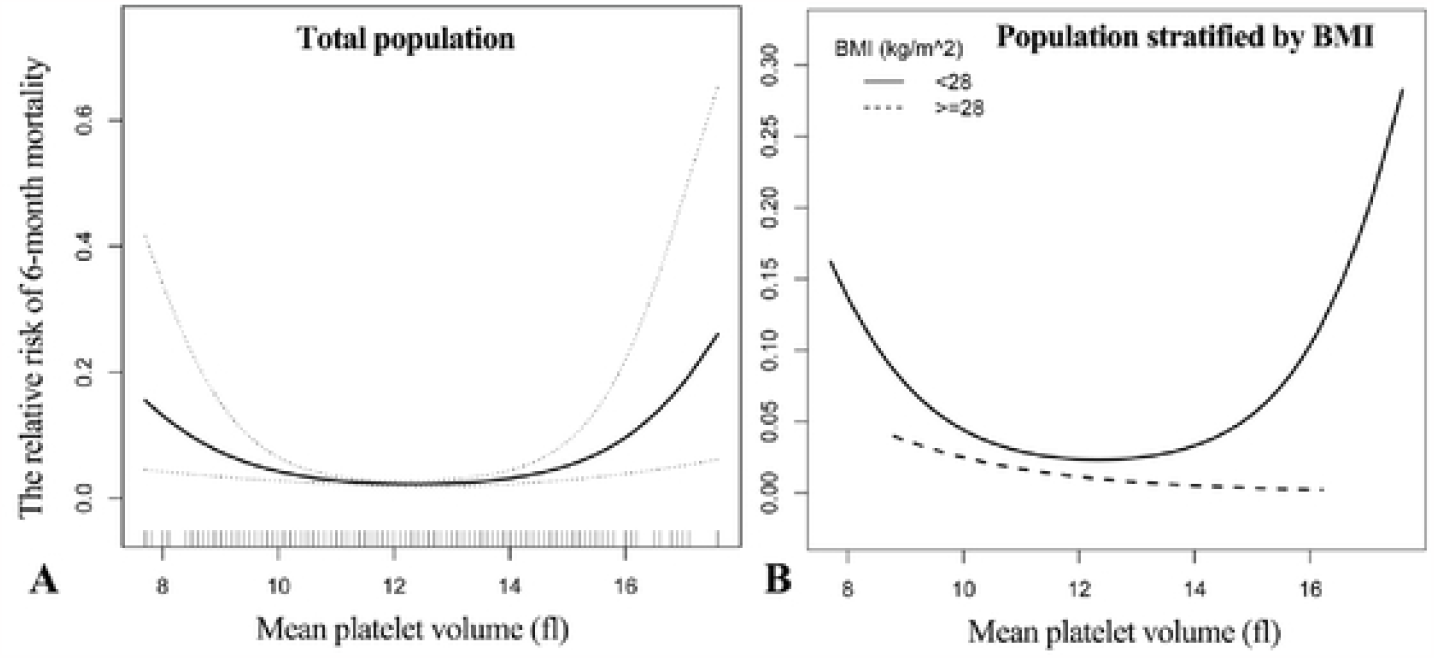

## Discussion

Our results imply that there is a U-shaped link between mean platelet volume and short-term mortality in HF in this cohort study. Both low and high MPV levels were linked to higher all-cause mortality in HF. However, this association did not hold true for obese individuals with HF. This discovery offers a priori support for upcoming model-driven studies and may have ramifications for how MPV levels are interpreted in clinical settings. As one of the most prevalent clinical markers, understanding the true relationship between MPV and a poor prognosis in HF will aid in the development of more accurate models. Our findings will also assist clinicians in making a more precise early evaluation of a patient’s future risk linked with MPV, even though this initial assessment is just approximate. Additionally, compared to other markers (blood glucose, potassium, etc.), which might alter as a result of various situations, MPV is a more stable measure.

Dilated chambers, poor contractility, local ventricular wall abnormalities, and combined atrial fibrillation may promote thromboembolism by facilitating intracardiac blood flow arrest, which is one of the possible mechanisms of poor prognosis in HF given that MPV is known to be a strong indicator of platelet activation and congestive HF is characterized by an increase in thromboembolic events(24). According to Jafri(25) et al., the coagulation system and platelet activation (increased plasma platelet factor 4, b-thromboglobulin) have been demonstrated to be more susceptible in patients with severe HF and poor ejection fraction, The relationship between platelet activation and a bad prognosis for HF stems from the possibility that these conditions may contribute to the development of the disease by producing ischemic episodes, impairing left ventricular function, and promoting intracardiac thrombosis. Chronic HF itself may increase the risk of platelet abnormalities and thrombosis through increased catecholamine release, haemodynamic changes, vascular factors, nitric oxide, cytokines, and comorbid conditions(26-28). The increased cytokine and catecholamine release observed in patients with severe HF is associated with platelet activation and higher levels of MPV. Therefore, by reducing excessive catecholaminergic activity, some studies have reduced MPV levels in patients with HF, thereby reducing mortality and hospitalization in HF(29, 30). In contrast, we discovered in our study that a low MPV was likewise linked to a higher chance of passing away. The occurrence is currently being reported for the first time, although no mechanisms have been found to explain it. Additionally, we discovered various MPV and mortality trends across states with obesity. We hypothesize that the absence of a correlation between the two in the obese group may be due to a lesser correlation between MPV and mortality in this cohort and a larger correlation between BMI and the risk of death. Of course, more data is required to support our findings given the tiny obese population in our study.

While there are some similarities and some discrepancies between our results and those of earlier studies(31, 32), they all point to the possibility of a higher MPV in HF. A previous study(19) first demonstrated that MPV levels above 9.1 fL were an independent predictor of short-to mid-term HF-related hospitalization in outpatients with stable HFrEF in SR. However, that study had a small sample size and was restricted to HF patients with reduced ejection fraction, whereas the current study was not. Kandis et al(2). conducted research. The best cutoff for indicating elevated risk was > 10.5 fl. However, the cohort of patients in that study showed higher levels of urea, serum creatinine, and systolic blood pressure in patients who passed away during follow-up, suggesting a stronger impact of the patient’s initial systolic blood pressure, BUN levels, and creatinine. In a cohort study(33) of the condition, an MPV value of > 9 fL was discovered to be a potential predictor of rehospitalization and 1-year mortality in patients with decompensated chronic HF in a cohort study. However, the study’s participants were those with decompensated chronic HF who were hospitalized (NYHA class IV HF or acute pulmonary oedema). The study has a limited sample size (n = 130) and no sensitivity analysis. By comparing the association between the HF marker BNP and MPV, Budak et al. **Error! Reference source not found**. discovered that MPV levels and brain natriuretic peptide (BNP) values were positively associated in patients referred to the emergency room with acute HF. According to the scientists, MPV can be a sign of the severity and clinical condition of acute HF disease. Taking into account study design, demographics, and other characteristics, we think the following factors are responsible for the results’ variations.

Our clinical relevance can be seen in the following ways: The previous finding that high MPV is strongly associated with a high risk of death in HF patients also holds true for the Chinese population, according to our findings. In addition, we discovered that lower MPV is also strongly associated with a high risk of death in HF patients, a finding that has never been previously reported. This study suggests that in the future, doctors utilizing MPV as an assessment indicator may need to include a patient’s obesity status when making clinical judgments or first prognosis assessments of patients. This offers fresh proof of MPV’s clinical value.

### Study advantages and limitations

We present a study with significant benefits, outlined below. Firstly, the use of large sample sizes enhances our statistical power. Secondly, the covariate information in our data is highly complete with few gaps. Thirdly, our work delves into non-linearity, providing further justification and insight. In comparison to previous studies, this allows for a greater understanding of the relationship between MPV and all-cause mortality. Lastly, we applied rigorous statistical corrections to address potential confounding variables, given the observational nature of our study.

However, some limitations should be acknowledged. Firstly, we only utilized baseline MPV measurements upon admission and were therefore unable to track MPV trends throughout hospitalization and their impact on outcomes. Secondly, the study was geographically and racially restricted to the Chinese population. Lastly, despite accounting for known potential confounders, residual confounders may still exist in any observational study.

### Conclusion

Our study investigated the association between baseline mean platelet volume and 6-month all-cause mortality in patients with heart failure. We found a U-shaped association, where the all-cause death rate increased at both low and high mean platelet volumes. However, we did not observe this connection in the population with obesity-related heart failure. These results have important clinical and public health implications, but additional prospective trials are needed to validate our findings and clarify their clinical implications.

## Data Availability

No

## Data Availability Statement

Publicly available datasets were analyzed in this study. This data can be found here: https://doi.org/10.13026/8a9e-w734.

## Ethics Statement

Studies involving human subjects were reviewed and approved by the Ethics Committee of the Fourth People’s Hospital of Zigong. In accordance with national legislation and institutional requirements, written informed consent for participation in this study was not required.

## Competing interests

The authors declare that the research was conducted in the absence of any commercial or financial relationships that could be construed as a potential conflict of interest. Funding

## Funding

This study was funded by the Science and Technology Support Plan of Guizhou Provincial Department of Science and Technology in 2020 (Project No. Qianke Support [2021] General 064).

Acknowledgments

We thank Chen Chi for their guidance and review of this manuscript.

## Authors’ Contributions

Y-LH collected documents and wrote the manuscript. QZ also contributed equally to this work. Y-LH and QZ are co-first authors. TZ reviewed and performed critical revision of the manuscript. All authors have read and agreed to the published version of the manuscript.

## Notes

### Competing Interest Statement

The authors have declared no competing interest.

### Clinical Trial

N

### Author Declarations

Ethics Committee of the Fourth People's Hospital of Zigong

